# The prevalence and developmental course of auditory processing differences in autistic children

**DOI:** 10.1101/2022.10.26.22280496

**Authors:** Bonnie K. Lau, Katherine A. Emmons, Adrian K.C. Lee, Jeff Munson, Stephen R. Dager, Annette M. Estes

## Abstract

Auditory processing differences, including hyper- or hyposensitivity to sound, aversions to sound, and difficulty listening under noisy, real-world conditions, are commonly reported in autistic individuals. However, the developmental course and functional impact of these auditory processing differences are unclear. In this study, we investigate the prevalence, developmental trajectory, and functional impact of reported auditory processing differences in autistic children throughout childhood using a longitudinal study design. Auditory processing differences were measured using the Short Sensory Profile, a caregiver questionnaire, in addition to adaptive behaviors and disruptive/concerning behaviors at 3, 6, and 9 years of age. Our results showed that auditory processing differences were reported in greater than 70% of the autistic children in our sample at all three timepoints, maintained a high prevalence through 9 years of age, and were associated with increased disruptive/concerning behaviors and difficulty with adaptive behaviors. Furthermore, in our sample of children, auditory processing differences at age 3 years predicted disruptive/concerning behaviors, difficulty with adaptive behaviors, and autism symptom severity at age 9 years. These findings warrant further investigations of the potential benefit of incorporating measures of auditory processing during routine clinical evaluations as well as interventions targeting auditory processing differences in autistic children.

**LAY SUMMARY:** We followed a cohort of autistic children longitudinally at 3, 6, and 9 years of age and found that more than 70% of the children had reported hyper- or hypo-sensitivity to sound, sound aversions, or difficulty listening in noisy environments at all three ages. These auditory processing differences were associated with increased hyperactivity and agitation, autism symptom severity, as well as difficulty with daily living skills, suggesting that auditory processing differences should be considered during routine clinical evaluations.

## INTRODUCTION

Autism spectrum disorder (ASD) is a neurodevelopmental disability characterized by difficulties with social communication and interaction, as well as restricted interests and repetitive behaviors, according to the Diagnostic and Statistical Manual of Mental Disorders, Fifth Edition (American Psychiatric Association, 2013). Characteristics of ASD present early in development and affect a range of behaviors including reduced eye contact, differences in verbal and nonverbal communication, repetitive motor movements, and heightened or reduced sensitivity to sensory input. In the United States, 1 in 44 children are estimated to have ASD, with boys four times more likely to be diagnosed than girls (Maenner, 2021). Early behavioral intervention has been shown effective in improving cognitive and behavioral outcomes in children with ASD, highlighting the importance of early screening and detection (Dawson et al., 2010; Klintwall, Eldevik, & Eikeseth, 2015; Vietze & Lax, 2020).

Differences in sensory processing have long been reported in autistic adults and children. Specific differences may include seeking out or avoiding visual stimulation, increased sensitivity to loud noises, and avoidance of certain sounds and textures (e.g., Leekam, Nieto, Libby, Wing, & Gould, 2007; O’Connor, 2012; Thye, Bednarz, Herringshaw, Sartin, & Kana, 2018). Under the current diagnostic criteria, sensory processing differences are a core feature of ASD, related to restrictive and repetitive behaviors. Atypical responses to sensory stimuli are common, with estimates of prevalence ranging from 70% - 90% (Baranek, David, Poe, Stone, & Watson, 2006; Kirby et al., 2022; Klintwall et al., 2011; Leekam et al., 2007; Tomchek & Dunn, 2007), and occur across all ages, levels of symptom severity, and sensory modalities (Ben-Sasson et al., 2009; Kern, 2006; O’Donnell, Deitz, Kartin, Nalty, & Dawson, 2012; Tomchek & Dunn, 2007; Tomchek, Huebner, & Dunn, 2014). While the prevalence of sensory processing differences in autism is well established, the developmental trajectory of sensory processing differences is less clear. Research on the topic has varied; some studies show decreases in sensory symptoms across childhood (Cheung & Siu, 2009), while others show no changes in symptoms (Green, Ben-Sasson, Soto, & Carter, 2012; McCormick, Hepburn, Young, & Rogers, 2016; Perez Repetto, Jasmin, Fombonne, Gisel, & Couture, 2017). The present study aims to elucidate the development of sensory processing differences in individual autistic children between the ages of 3 and 9 years.

Successful integration of sensory input is crucial for daily functioning. In autistic adults and children, sensory processing challenges contribute to differences in social interaction and communication and effect daily functioning and academic performance (Dellapiazza et al., 2018; Howe & Stagg, 2016; Kojovic, Ben Hadid, Franchini, & Schaer, 2019; Suarez, 2012; Thye et al., 2018). Sensory processing differences are also associated with increased behavioral and emotional difficulties (Baker, Lane, Angley, & Young, 2008), lower adaptive functioning (Kojovic et al., 2019), increased social impairments (Hilton et al., 2010; Janet K. Kern et al., 2007), and parental stress (Ben-Sasson, Soto, Martínez-Pedraza, & Carter, 2013). In young autistic children, early sensory dysregulation may hinder the development of joint attention, language, and social play (Baranek et al., 2013; Miller Kuhaneck & Britner, 2013). A growing body of research suggests atypical responses to sensory input may precede differences in social interaction and communication, wherein early neural dysfunction produces cascading effects on social development (Baranek et al., 2018; Damiano-Goodwin et al., 2018). Infants who go on to develop ASD show increased sensory seeking compared to those who do not, leading to reduced social orienting and increased social symptom severity (Baranek et al., 2018). Thus, early screening for sensory processing differences may play a role in early detection of ASD.

Compared with other sensory modalities, auditory processing differences are frequently reported in autistic adults and children. Commonly reported auditory symptoms include hyper- or hyposensitivity to sound, aversions or unusual interest in specific sounds, and difficulty listening in noisy environments (see O’Connor, 2012 for review). Furthermore, some evidence suggests behavioral differences are more closely linked to auditory processing differences compared to other sensory domains (Germani et al., 2014).

Prior behavioral and neurophysiological research suggests alterations in auditory processing may be an early marker for ASD (Germani et al., 2014; Miron et al., 2021; Riva et al., 2018). In a recent study, parent reports of auditory processing differences at 24 months of age were associated with subsequent ASD diagnoses at age three years in infants with familial likelihood for ASD (Germani et al., 2014). Similarly, abnormalities in auditory event-related potentials recorded by electroencephalography (EEG) at age 12 months were associated with expressive vocabulary and critical M-CHAT items at age 20 months in infants at high likelihood of developing ASD (Riva et al., 2018). Based on the retrospective analysis of newborn hearing screening data, infants who were later diagnosed with ASD had longer auditory brainstem response latencies compared to infants who did not go on to develop ASD, suggesting neurophysiological variation may be present at birth in infants who go on to develop ASD (Miron et al., 2021).

Here, we aim to investigate the prevalence, developmental trajectory, and functional impacts of reported auditory processing differences in autistic children using a longitudinal study design. The Short Sensory Profile (SSP; McIntosh, Miller, Shyu, & Dunn, 1999), a widely used caregiver questionnaire, was chosen as a measure of sensory processing differences in our sample. Functional behaviors were assessed using the Vineland Adaptive Behavior Scales (VABS; Sparrow & Cicchetti, 1989) and the Aberrant Behavior Checklist (ABC; Aman & Singh, 1986). The research questions under investigation were:

1. What is the prevalence of reported auditory processing differences in this longitudinal sample?
2. What is the developmental course of auditory processing differences across childhood, from age 3 years to 9 years?
3. What are the functional consequences of auditory processing differences in autistic children?

## METHODS

### Participants

Data from a larger longitudinal study on the neurobiology and developmental course of ASD conducted at an autism center at a major university, was retrospectively analyzed. The full study sample consisted of 74 children who were diagnosed with ASD at age 3, with data obtained between 1990s and 2000s for this initial time point. This cohort of participants were then followed at 6 and 9 years of age. Children who had a history of sensory or motor impairment including hearing loss, traumatic brain injury, major physical anomalies, genetic disorders associated with ASD such as Fragile X, or other neurological impairments were excluded (see Dawson et al., 2004 for further details). An initial research diagnosis according to the DSM-IV (American Psychiatric Association, 1998) criteria was established at 3 years of age by a licensed clinical psychologist or a qualified graduate student in clinical psychology using: (1) the Autism Diagnostic Interview-Revised (ADI-R; Rutter, LeCouteur, & Lord, 2003), a standardized parent interview; (2) the Autism Diagnostic Observation Schedule-Generic (ADOS-G; Lord, Rutter, DiLavore, & Risi, 2003), a standardized semi-structured play observation, scored with the revised algorithm (Gotham, Risi, Pickles, & Lord, 2007); (3) medical and family history; (4) cognitive test scores; and (5) clinical observation and judgment. This diagnostic battery was repeated at 6 years and 9 years of age. All children with data on the SSP at one or more time points were included in this analysis: 49 children had SSP data at 3 years, 29 at 6 years, and 52 at 9 years of age. SSP data was obtained from 20 children at one time point, 34 children at two time points, and 14 children at all three time points. Written informed consent was obtained from a parent or legal representative prior to participation in this study. Ethical approval was given for this study by the Institutional Review Board at the University of Washington and all analyses were conducted in accordance with the protocols approved.

### Procedure

Reported sensory processing differences, adaptive behaviors, disruptive/concerning behaviors, cognition, and vocabulary were measured at 3, 6 and 9 years of age. Descriptive data for the study sample are provided in Table 1.

**Table 1.** Descriptive data for study sample – Mean (SD)

|  | 3 Years | 6 Years | 9 years |
| --- | --- | --- | --- |
| Sensory Processing | *N*=49 | *N*=29 | *N*=52 |
| SSP Total Score | 159.67(23.17) | 153.58 (29.36) | 155.98(25.32) |
| Auditory Filtering | 19.23(4.32) | 20.06(3.75) | 19.00(4.12) |
| Symptom Severity | *N*=49 | *N*=29 | *N*=52 |
| ADOS severity score | 6.59(1.67) | 7.28(1.87) | 7.50(2.11) |
| Adaptive Behavior | *N*=49 |  | *N*=52 |
| VABS-C | 60.32(9.40) |  | 54.10(20.17) |
| Disruptive/Concerning | *N*=49 | *N*=27 | *N*=52 |
| Behavior | 10.92(7.93) | 10.26(7.36) | 9.50(9.52) |
| ABC-AG | 15.45(9.82) | 13.81(8.11) | 12.96(10.08) |
| ABC-HYP |  |  |  |
| Cognition | *N*=47 | *N*=29 | *N*=50 |
| MSEL-C | 58.83(15.31) |  |  |
| DAS-GCA |  | 65.52(21.08) | 79.30(24.68) |
| Expressive Vocabulary | *N*=45 | *N*=23 | *N*=31 |
| CDI – Produced | 10.76(9.84) |  |  |
| EVT |  | 72.04(24.41) | 88.74(15.08) |
| Receptive Vocabulary | *N*=45 | *N*=24 | *N*=46 |
| CDI – Pnderstood | 36.78(51.93) |  |  |
| PPVT-III |  | 71.63(23.92) | 80.28(28.76) |

### Measures

#### Sensory Processing

Sensory processing was assessed using the SSP, a standardized 38-item caregiver questionnaire developed to identify children with sensory processing difficulties and associated behaviors (McIntosh et al., 1999). Each item on the SSP is rated on a Likert scale ranging from 1 to 5 with a score of 1 assigned to behaviors that are “always” occurring and a score of 5 assigned to behaviors that are “never” occurring. Thus, a lower score indicates a higher occurrence of sensory processing difficulties. The item raw scores are aggregated into 7 subscale scores: Tactile Sensitivity, Taste/Smell Sensitivity, Movement Sensitivity, Underresponsive/Seeks Sensation, Auditory Filtering, Low Energy/Weak, and Visual/Auditory Sensitivity. For each subscale score as well as the SSP Total score, cutoff points based on the normative sample are provided for classification into the categories of Typical Performance, Probable Difference (−1 to −2 SDs below the mean) and Definite Difference (>2 SDs below the mean). The normative scores were established from the full-length Sensory Profile (Dunn, 1999), which was standardized on 1,037 typically developing children.

To determine the prevalence of auditory processing differences in the study sample, the percentage of children with Auditory Filtering Subscale Scores in the Probable Difference and Definite Difference range was quantified. The prevalence of Auditory Filtering differences relative to other domains was determined by comparing across the 7 subscale scores. The six items in the Auditory Filtering subscale capture hyposensitivity to sound and difficulty attending to or tuning out sound in a noisy environment (items shown in Table 2).

**Table 2.** SSP Auditory Filtering Items

| Item | Subscale |
| --- | --- |
| 22. Is distracted or has trouble functioning if there is a lot of noise around | Auditory Filtering |
| 23. Appears to not hear what you say (e.g., does not 'tune in' to what you say, appears to ignore you) | Auditory Filtering |
| 24. Can't work with background noise (e.g., fan, refrigerator) | Auditory Filtering |
| 25. Has trouble completing tasks when the radio is on | Auditory Filtering |
| 26. Doesn't respond when name is called but you know the child's hearing is OK | Auditory Filtering |
| 27. Has difficulty paying attention | Auditory Filtering |

For participants with missing data on the SSP, the average of the remaining scores in the subscale was calculated, and this value replaced the missing score(s) in that subscale. This approach was selected to control for bias while still maintaining representation of the participant’s sensory profile. One SSP at 9 years was excluded because more than 20% of the items were incomplete.

#### Symptom Severity

The ADOS-G, a semi-structured standardized measure that assesses social interaction, communication, play, and repetitive behaviors was administered to all participants as part of the initial diagnostic confirmation process at 3 years of age. Symptom severity was determined with the ADOS-G Total Score, a sum of raw scores on diagnostic items which ranged from 1 to 10.

#### Assessment of Functional Behaviors

Personal and social functioning was assessed with the Vineland Adaptive Behavior Scales (VABS; Sparrow & Cicchetti, 1989), a standardized 297-item parent interview. The VABS assesses adaptive behaviors in four domains: communication, daily living skills, socialization, and motor, with norms derived from a sample of 4,800 individuals with and without disabilities. The VABS adaptive behavior composite (VABS-C) is calculated from the four domain scores. As only two of the four VABS domains were administered at age 6, the VABS-C was obtained only at 3 years and 9 years of age. Thus, analyses involving the VABS-C was not conducted at the 6-year time point.

Disruptive and concerning behaviors were measured with the Aberrant Behavior Checklist (ABC; Aman & Singh, 1986), a 58-item parent report measure. The items belong to five subscales: (1) Irritability, agitation, crying; (2) lethargy, social withdrawal; (3) stereotypic behavior, (4) hyperactivity, non-compliance; and (5) inappropriate speech. Each item is scored on a scale of 0-3 with 0 indicating “not at all a problem” and 3 indicating “the problem is severe in degree”. Thus, a higher score indicates a higher occurrence of disruptive/concerning behaviors. To investigate the effect of sensory processing on mood and behavior, the analyses in this study focus on the Agitation (ABC-AG; irritability, agitation, crying) and Hyperactivity (ABC-HYP; hyperactivity, noncompliance) subscales.

Intellectual ability was assessed with the Mullen Scales of Early Learning (MSEL; Mullen, 1995) at age 3. The Early Learning Composite (MSEL-C) is calculated from the Visual Reception, Receptive Language, Expressive Language and Fine Motor subscale T scores. At ages 6 and 9, cognition was assessed with the Differential Abilities Scale (DAS; Elliott, 1990), a battery of tests used to measure verbal, reasoning, perceptual, and memory skills. The School Age Level administered included six core subtests, yielding the General Conceptual Ability standard score (DAS-GCA).

Expressive and receptive vocabulary at age 3 was assessed with the MacArthur-Bates Communicate Development Inventories-Words and Gestures (CDI-WG), a parent report measure. At age 6 and 9, the Expressive Vocabulary Test (EVT; Williams, 1997) and the Peabody Picture Vocabulary Test – 3^rd^ Edition (PPVT-III; Dunn&Dunn, 1997) was administered with each child. The standard score for each measure is reported in Table 1.

## RESULTS

### Prevalence of reported auditory filtering differences across time

SSP scores were first analyzed to determine the prevalence of reported auditory processing differences in the study sample. Figure 1 shows the percentage of children with scores in the Definite or Probable Difference range across SSP subscales at each time point. A large proportion of children reported sensory processing differences, especially in the auditory domain. More than 70% of children scored within the Definite or Probable Difference range on the Auditory Filtering subscale at each time point; and the prevalence increased over time. These numbers were consistent with the overall high percentage of children in the sample that were rated as having some degree of sensory processing difference based on the SSP Total score: 76% at 3 and 6 years and 81% at 9 years. Auditory Filtering had the highest percentage of reported difference of any subscale at all three time points. However, at 3 years, the difference between the percentage of children with reported sensory processing differences in the Auditory Filtering subscale reached statistical significance only in comparison to the Low Energy/Weakness (*p*<0.001, fisher exact test) and the Movement Sensitivity subscale (*p*<0.001, fisher exact test; *p*>0.05 for all other subscales). At 6 years, the difference reached statistical significance in comparison to the Movement Sensitivity subscale only (*p*<0.001, fisher exact test; *p*>0.05 for all other subscales). By 9 years of age, the number of children with a reported difference in Auditory Filtering (80%) surpassed all other subscales (<60% for all others). This difference was statistically significant for the the Low Energy/Weakness (*p*<0.001, fisher exact test), Movement Sensitivity (*p*<0.001, fisher exact test), and the Visual/Auditory Sensitivity subscale (*p*=0.03, fisher exact test; *p*>0.05 for all other subscales).

**Figure 1.**
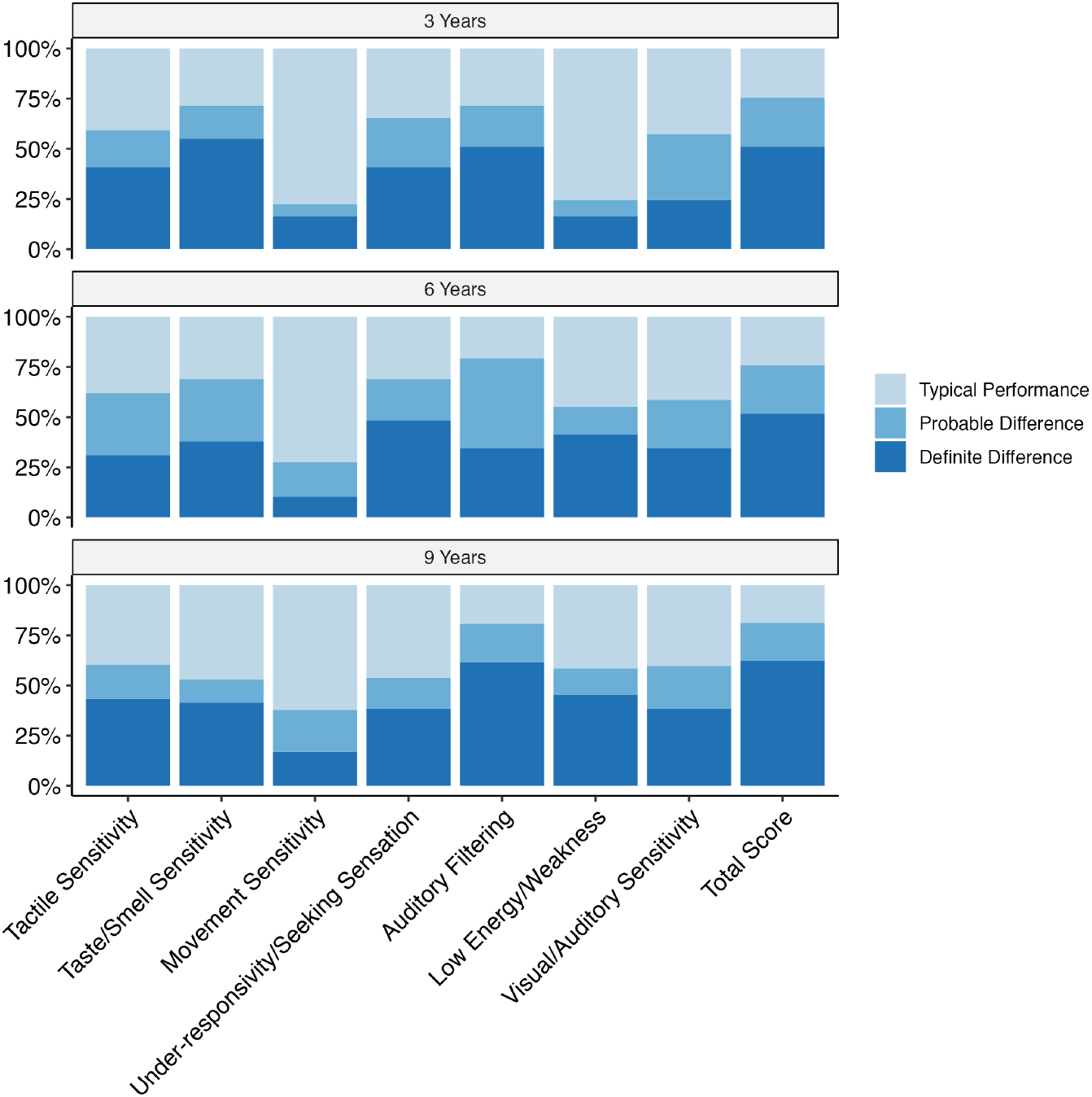
Percentage of children with reported sensory processing differences in the Definite Difference, Probable Difference, and Typical Performance range in each subscale of the SSP at 3, 6, and 9 years of age.

### Developmental course of reported auditory processing differences

To investigate the developmental course of reported auditory processing differences, change in the Auditory Filtering Subscale Score over time was assessed. For most subjects, the Auditory Filtering Subscale Score appeared to remain stable across the three time points (Fig. 2). A general linear mixed model with the SSP Auditory Filtering Subscale Score as the dependent variable, timepoint as a fixed effect, and a random intercept and a random slope for timepoint was fit according to the equation as follows (Eq.1):

**Figure 2.**
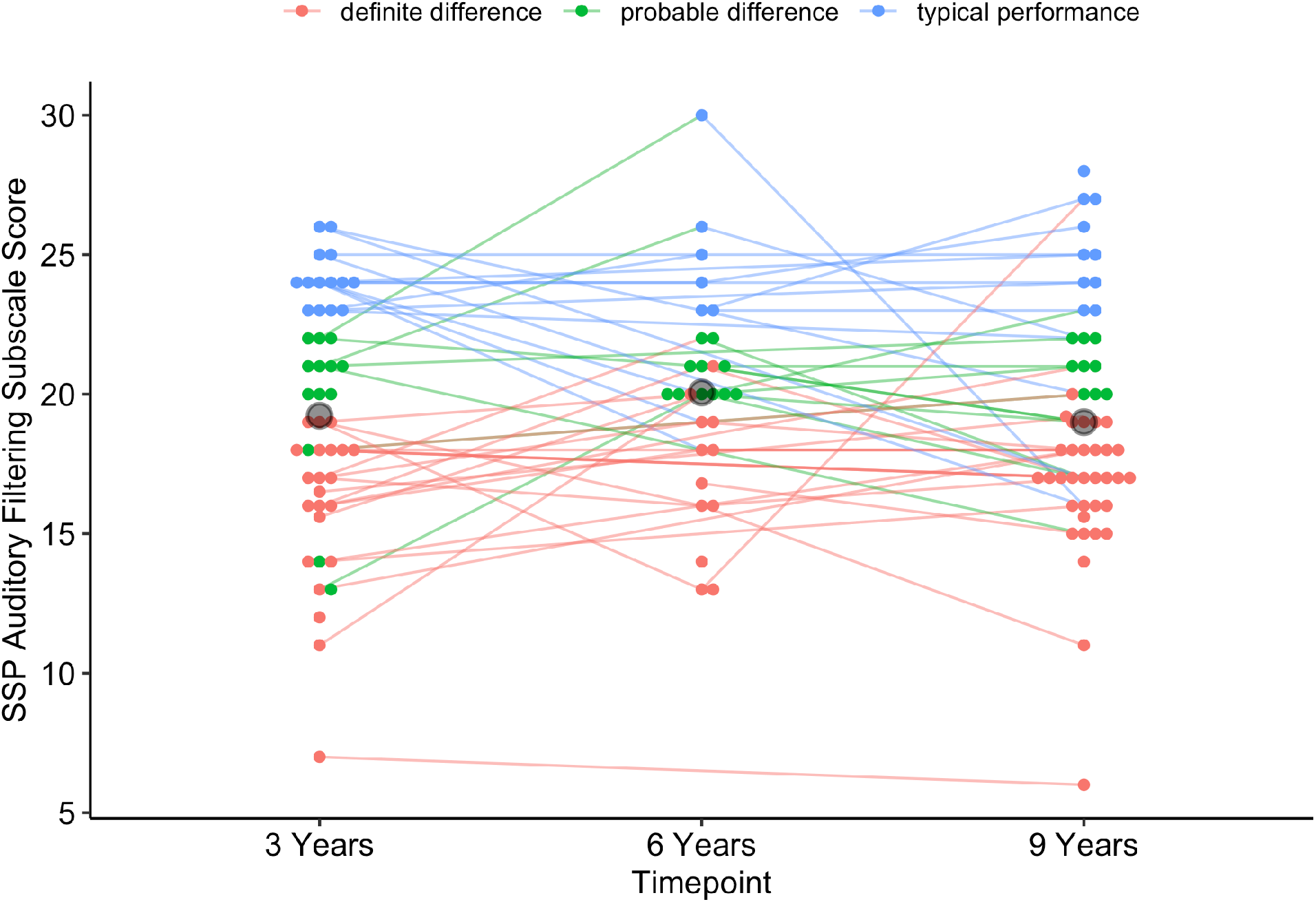
SSP Auditory Filtering Subscale Score as a function of time point. Individual participant trajectories are color-coded with red indicating definite difference, green indicating probable difference, and blue indicating typical performance. Large gray circle showing mean at each time point.

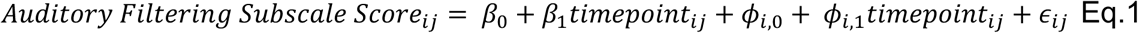

where *β*_1_ is the fixed effect of time point (3, 6, or 9 years), the primary effect of interest, and *ϵ*_*ij*_ is an error term (Table 3). This model was selected for its ability to handle children with missing data from one or more time points and its ability to capture individual variability across subjects: the random intercept accounts for variability in the Auditory Filtering Subscale Score while the random slope captures variability in how the score changes over time for each subject. The random effects were modeled with an autoregressive covariance structure, which considers correlation to be highest for adjacent time points. Results of the model revealed no significant fixed effect of time point (*p*=0.70) confirming that SSP Auditory Filtering Subscale Score remained stable from 3 to 9 years of age.

**Table 3.** Linear mixed effect model output for change in SSP Auditory Filtering Subscale Score over time

| Fixed Effects | $\beta$ Estimate | 95% Confidence Intervals | p-value |
| --- | --- | --- | --- |
| *Base Model – Fixed effect of time point, random slope and intercept* |  |  |  |
| Intercept | 19.55 | 17.98, 21.12 | <0.001 |
| Time point | -0.14 | -0.88, 0.59 | 0.70 |
| *Model controlling for cognition* |  |  |  |
| Intercept | 19.24 | 16.68, 21.80 | <0.001 |
| Time point | -0.08 | -0.93, 0.77 | 0.84 |
| Intellectual Ability | 0.002 | -0.04, 0.04 | 0.89 |
| *Model controlling for ADOS severity score* |  |  |  |
| Intercept | 21.85 | 19.03, 24.67 | <0.001 |
| Time point | 0.03 | -0.72, 0.78 | 0.93 |
| ADOS severity | -0.37 | -0.76, 0.01 | 0.056 |

During secondary analyses, two additional models were run to control for intellectual ability and symptom severity. In the first model, standard scores for the cognitive assessments administered at each time point were entered into the base model as a covariate. The model results revealed that intellectual ability was not a significant covariate (*p*=0.89). After adjusting for intellectual ability, the parameter estimate for time point shifted, although the fixed effect of time point remained non-significant (*p*=0.84). In the second model, the symptom severity score from the ADOS obtained at each time point was entered as a covariate. Model results revealed that the ADOS Severity Score was a not a significant covariate (*p*=0.06). Again, the parameter estimate for time point was shifted after controlling for the ADOS Severity Score but the fixed effect of time point did not reach significance *(p*=0.94). Results of these additional models provide further evidence that even after controlling for intellectual ability and symptom severity, the Auditory Filtering Subscale Score is comparable at all three time points.

Finally, to compare the developmental course of auditory processing to overall sensory processing differences, change in the SSP Total score over time was assessed using a general linear mixed model with the SSP Total score as the dependent variable, timepoint as a fixed effect, and a random intercept and a random slope for timepoint. Results of the model revealed no significant fixed effect of time point (*p*=0.38) showing that SSP Total score also remained stable from 3 to 9 years of age.

### Relationship between auditory processing and functional behaviors

The association between the Auditory Filtering Subscale Score and disruptive/concerning behavior, adaptive behavior, symptom severity, cognition, and vocabulary scores at 3 and 9 years of age was assessed using a Pearson Correlation coefficient (Table 4). This analysis was not conducted at age 6 due to missing data; SSP data was obtained from only 29 children and the VABS-C was not obtained at this time point.

**Table 4.** Correlations between SSP Auditory Filtering Score and functional behaviors

|  | 3 Years<br>r (N) | 9 years<br>r (N) |
| --- | --- | --- |
| Adaptive Behavior<br>VABS-C | 0.35 (44)** | 0.35 (50)** |
| Disruptive/Concerning<br>Behaviors | -0.56 (49)** | -0.40 (52)** |
| ABC-AG | -0.68 (49)** | -0.51 (52)** |
| ABC-HYP |  |  |
| Symptom Severity<br>ADOS severity score | -0.37 (49)** | -0.13 (52) |
| Intellectual Ability | 0.16 (47) | 0.001 (50) |
| Vocabulary |  |  |
| Expressive | 0.15 (45) | 0.18 (31) |
| Receptive | -0.10 (45) | 0.58 (44) |

Pearson Correlations, \* $p < 0.05$ , \*\* $p < 0.01$ level (2-tailed)

At age 3, there was a highly significant correlation between the Auditory Filtering Subscale Score and the ABC-HYP, the ABC-AG, the VABS-C and the ADOS severity score (Fig. 3A, *p*<0.02 in all cases). The association between the Auditory Filtering Subscale Score and the two ABC scores was strong and negative, indicating that more reported auditory processing differences was correlated with increased agitation and hyperactivity. There was also a moderate association between the Auditory Filtering Subscale Score and symptom severity as well as adaptive behaviors, with more reported auditory processing differences correlated to higher symptom severity and increased difficulty with adaptive behaviors.

**Figure 3.**
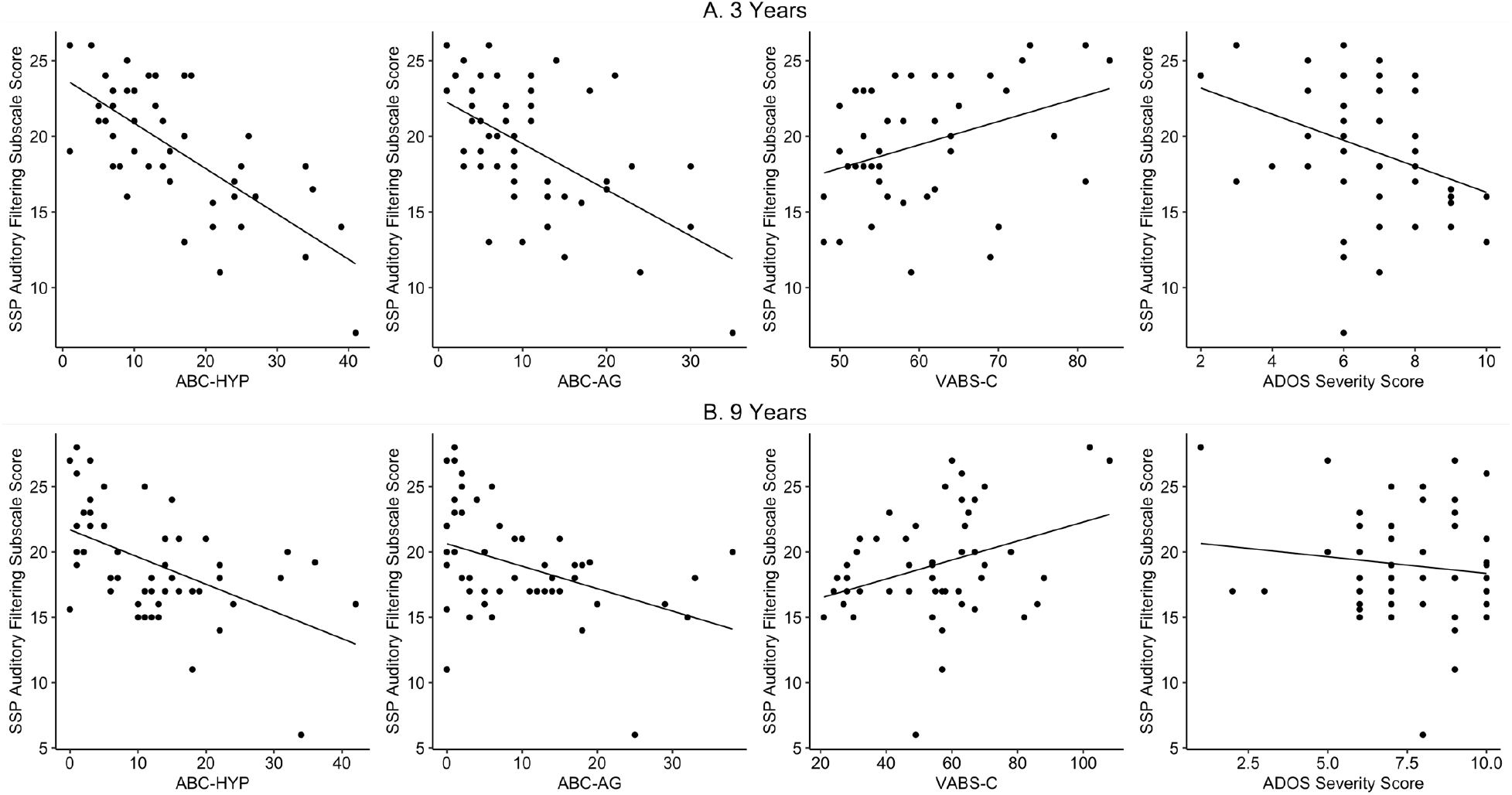
**A)** Relationship between SSP Auditory Filtering Subscale Score and ABC-HYP, ABC-AG, VABS-C, and ADOS Severity Score at age 3 years. **B)** Relationship between SSP Auditory Filtering Subscale Score and ABC-HYP, ABC-AG, VABS-C, and ADOS Severity Score at age 9 years.

By age 9, similar patterns of association were observed with a highly significant correlation between the ABC-HYP, the ABC-AG, and the VABC-C (Fig. 3B, *p*<0.01 in all cases). Thus, at both time points, reported auditory processing differences were associated with increases in disruptive/concerning behaviors and difficulty with adaptive behaviors. However, the association with symptom severity by 9 years of age was no longer significant (Fig. 3B, *p*=0.256). At both time points, no correlation between reported auditory processing differences, intellectual ability, or vocabulary scores was observed (*p*>0.05 in all cases).

The predictive relationship of reported auditory processing differences at 3 years with agitation, hyperactivity, adaptive behaviors, and symptom severity at 9 years was further assessed using linear regression. Four separate linear regression models were run with the Auditory Filtering Subscale Score as the predictor variable (Figure 4). The model results indicated that the Auditory Filtering Subscale Score at age 3 was a significant predictor of all four measures at age 9 (Table 5). For disruptive/concerning behaviors, the Auditory Filtering Subscale Score at age 3 explained 14.5 % of the variance in the ABC-AG scores and 23.7% of the variance in the ABC-HYP scores at age 9. A one-point increase in the Auditory Filtering Subscale Score resulted in a 0.8 decrease in ABC-AG and a 1.15 decrease in the ABC-HYP scores. For adaptive behaviors, the Auditory Filtering Subscale Score at age 3 explained 7% of the variance of the VABS-C score at age 9 and a one-point increase in the Auditory Filtering Subscale Score resulted in a 1.1 improvement in VABS-C score. For symptom severity, the Auditory Filtering Subscale Score at age 3 score explained 2.4% of the variance in the ADOS severity score at age 9 and a one-point increase in the Auditory Filtering Subscale Score was associated with a 0.083-point decrease in ADOS severity score.

**Table 5.** SSP Auditory Filtering Subscale Score at 3 years predicts functional behaviors at 9 years

|  | Coefficient | 95% Confidence<br>Intervals | *p*-value | R<sup>2</sup> |
| --- | --- | --- | --- | --- |
| ABC-AG | -0.80 | -1.14 – -0.47 | <0.001 | 0.15 |
| ABC-HYP | -1.15 | -1.51 – -0.79 | <0.001 | 0.24 |
| VABS-C | 1.10 | 0.35 – 1.85 | 0.005 | 0.08 |
| ADOS severity | -0.08 | -0.16 – -0.002 | 0.05 | 0.03 |

**Figure 4.**
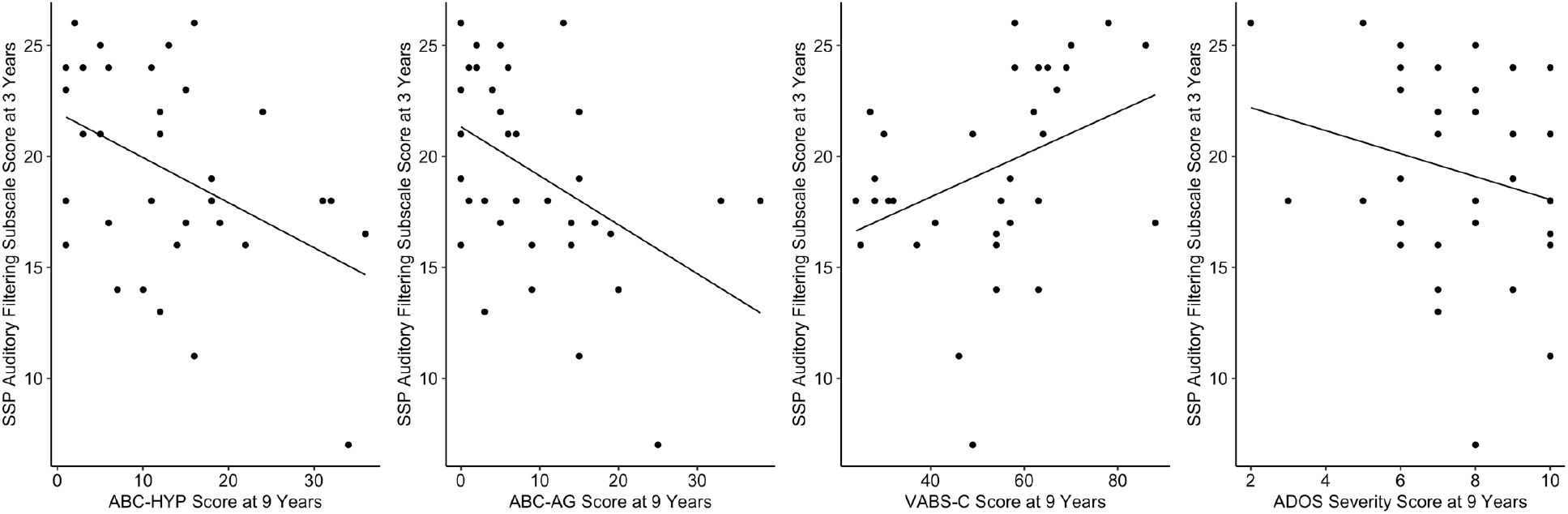
Relationship between SSP Auditory Filtering Subscale Score at 3 years and ABC-HYP, ABC-AC, VABS-C and ADOS Severity Score at 9 years.

## DISCUSSION

Results from this study reveal three main findings. First, a large proportion (>70%) of children in our sample had reported auditory processing differences at each time point. This finding is consistent with Tomchek & Dunn (2007) who observed differences in the Auditory Filtering subscale of the SSP in over 75% of their sample of autistic children aged 3 to 6 years. Second, auditory processing differences remained stable and elevated from age 3 to 9 years, even after controlling for intellectual ability and autism symptom severity. Again, this result is consistent with the literature. For example, McCormick et al. (2016) demonstrated no significant change in SSP total scores or subscale scores in autistic children between ages 2 and 8 years. Finally, auditory processing differences at ages 3 and 9 years were associated with increased disruptive/concerning behaviors and difficulty with adaptive behaviors. This finding is consistent with prior research showing associations between sensory processing differences and increased disruptive/concerning behaviors and difficulty with adaptive behaviors in autistic adults and children (Baker et al., 2008, 2008; Hilton et al., 2010; Janet K. Kern et al., 2007; O’Donnell et al., 2012).

Several novel findings also emerged from our analyses. The number of children with reported auditory processing differences at age 9 years surpassed all other subscales of the SSP. In other words, parents reported auditory processing differences more than any of the other domain of the SSP at age 9 years. In a cross-sectional study of young autistic children and adults aged 32 months to 38 years, Leekam et al. (2007) found that auditory symptoms were consistent between age groups, while symptoms in other domains (e.g., oral) were less common in older participants. Thus, our findings expand on the current literature by demonstrating that auditory processing differences are stable and elevated not just across age groups, but within the same individual during early childhood development. It may also be the case that auditory processing challenges become more apparent during school-age, as children transition from being at home to spending more time in noisy classroom environments. Thus, these findings may have important implications for caregivers, educators, and service providers, as autistic children may need additional supports to cope with auditory processing differences during school age.

Also significant was the finding that auditory processing differences at age 3 years predicted disruptive/concerning behaviors, difficulty with adaptive behaviors, and autism symptom severity at age 9 years. That is, auditory processing differences in early childhood were correlated with poorer outcomes in later childhood. This finding underscores the importance of comprehensive clinical evaluations that include measures of auditory processing function in autistic children. Moreover, it suggests interventions targeting auditory processing differences in early childhood may be particularly beneficial for future adaptive functioning. Future studies should evaluate whether auditory processing interventions improve outcomes for young autistic children.

There are several limitations to the present study. Namely, this study focused on caregiver-reported differences rather than physiological or behavioral measure of auditory alterations. Direct measure of sensory processing may provide more objective data and could help reveal mechanisms related to sensory alterations. While many aspects of the underlying neurophysiology of sensory processing alterations in autism is not well understood, there is some emerging evidence to suggest a common neurophysiological basis for restrictive and repetitive behaviors and structural properties of cerebellar and corpus collosum white matter in infants who go on to develop ASD (Wolff et al., 2017). Recent research has also begun to identify associations between basic auditory processing and severity of autism symptoms (Brandwein et al., 2014). Future research should aim to elucidate the neurophysiological underpinnings of sensory processing differences and how they may relate to early brain changes associated with ASD.

In conclusion, results from this study demonstrate 1) auditory processing differences are common in autistic children aged 3 to 9 years, 2) for many children, auditory processing differences are stable or increase from 3 to 9 years of age, 3) reported auditory processing differences at ages 3 and 9 years are associated with increased disruptive/concerning behaviors and difficulty with adaptive functioning. Auditory processing differences at age 3 years predicted disruptive/concerning behaviors, difficulty with adaptive behaviors, and autism symptom severity at age 9 years. These findings suggest the potential benefit of incorporating measures of auditory processing during clinical evaluations conducted in autistic children and developing interventions targeting auditory processing differences in early childhood.

## Data Availability

All data produced in the present study are available upon reasonable request to the authors

## ACKNOWLEDGEMENTS

We thank the participants and their families for their participation in this research.

